# Insights into a Rare Clinical Phenomenon: Isolated Native Valve Endocarditis After Prosthetic Valve Implantation

**DOI:** 10.1101/2025.07.12.25331439

**Authors:** Sujoy Khasnavis, Majd Alhuarrat, Caroline Delbourgo Patton, Shaunak Mangeshkar, Robert Faillace, Michael Grushko

**Affiliations:** Jacobi-North Central Bronx Hospital and Medical Center, Bronx NY; D. Samuel Gottesman Library, Albert Einstein College of Medicine, Bronx, NY

**Keywords:** Isolated native valve endocarditis, transcatheter aortic valve implantation, prosthetic valve implantation, mitral endocarditis, right heart endocarditis

## Abstract

**Introduction:** Prosthetic valve endocarditis (PVE) is a known complication of prosthetic valve implantation (PVI). The incidence of selective native valve endocarditis without concurrent PVE, or isolated native valve endocarditis (INVE), is much lower and largely unheard of in literature.

**Methods:** We examine here the specific circumstances in which INVE manifests as well as management and complications of this rare form of endocarditis. A systematic review of four major databases was carried out to identify INVE and PVE cases after various PVIs. INVE was compared to PVE across categories such as patient demographics, method and type of PVI prior to endocarditis, pathogen type, management, and outcomes.

**Results:** INVE comprised 18.2 % of valvular endocarditis cases after transcatheter aortic valve implantation (TAVI) and 6.5 % of valvular endocarditis cases after surgical aortic valve implantation (SAVI). In the TAVI group, mean age of the INVE cohort was 79.5 +− 5 years and it was observed more frequently among males. Prior to most INVE cases, TAVI had been performed by transfemoral route. INVE was more closely linked to diabetes (DM) and chronic obstructive pulmonary disease (COPD) than TAVIE (transcatheter aortic valve implant endocarditis). INVE was less closely linked to chronic kidney disease (CKD) than TAVIE. Gram positive infection rates were similar between the groups. Surgery was done less frequently after INVE than TAVIE. Compared to TAVIE, INVE mortalities were more likely to occur at follow up and were linked to heart failure, renal failure, and septic shock. Total long term mortalities were similar between the two groups.

**Conclusion:** INVE makes up a measurable proportion of endocarditis cases after TAVI and shares characteristics, predictors, pathogens, and outcomes linked to TAVIE. Compared to TAVIE, INVE is more closely linked to DM and COPD and less to CKD. It manifests more on mitral valves than right heart valves and presents higher rates of delayed mortality. Given INVE’s long term mortality and ability to manifest on different valves, thorough cardiac evaluation and close long term follow up should be considered if endocarditis is suspected after TAVI. Non-mitral INVE and surgical implant based INVE are less frequently reported on and warrant further investigation.

**Key Points:** *Question:* What are the incidence, associations, predictors, implicated causes, and outcomes of selectiveand isolated native valve endocarditis following prosthetic valve implantation?

*Findings:* This systematic review found that isolated native valve endocarditis was reported most frequently after transcatheter aortic valve implants and particularly in mitral valves. Infrequently, isolated native endocarditis has also been reported in right heart valves and after surgical aortic valve implants. Diabetes and pulmonary disease are associated with isolated native endocarditis after transcatheter implants.

*Meaning:* The development of isolated native mitral valve endocarditis should be closely screened for in diabetic and pulmonary patients with transcatheter aortic valve implants. This phenomenon needs further study in non-aortic, non-transcatheter, and non-mitral endocarditis populations. Moreover, patients with this rare endocarditis require close long term follow up as mortalities after hospitalization are significantly higher than in hospital.

## Introduction

Prosthetic valve endocarditis (PVE) is an established complication of mechanical and bioprosthetic valve placement^1,2^. Generally, infective endocarditis (IE) has a predilection for prosthetic valves over native valves given the nidus that the prostheses provide for bacterial attachment and infection^3,4^. PVE also carries a higher mortality rate than does native valve endocarditis (NVE). The incidence of NVE subsequent to PVE has also been described and is known to carry a poorer prognosis than isolated PVE^4,8^. In the setting of PVI, the development of isolated native valve endocarditis (INVE) without PVE is an unusual phenomenon. Select retrospective reviews have described instances of mitral INVE after PVIs^1,3,6,9,12^. Case reports have also described instances of mitral and aortic INVE after atypical bacterial infections^18,19,20^. A comprehensive review detailing the specifics of INVE has not been performed in prior studies however. This review provides a summary of the literature on INVE after various PVIs including transcatheter valve implantation and surgical valve implantation with the purpose of understanding the patient characteristics, potential contributors, management strategies, and outcomes associated with this rare and unusual type of IE.

## Methods

### Search strategy

A systematic review of studies reporting INVE was performed across databases of Pubmed, Embase, Cochrane Library, and Web of Science. Search terms encompassed PVI types including transcatheter valve implants and surgical valve implants as well as PVE and NVE. Table 1 shows the complete search criteria.

**Table 1.**
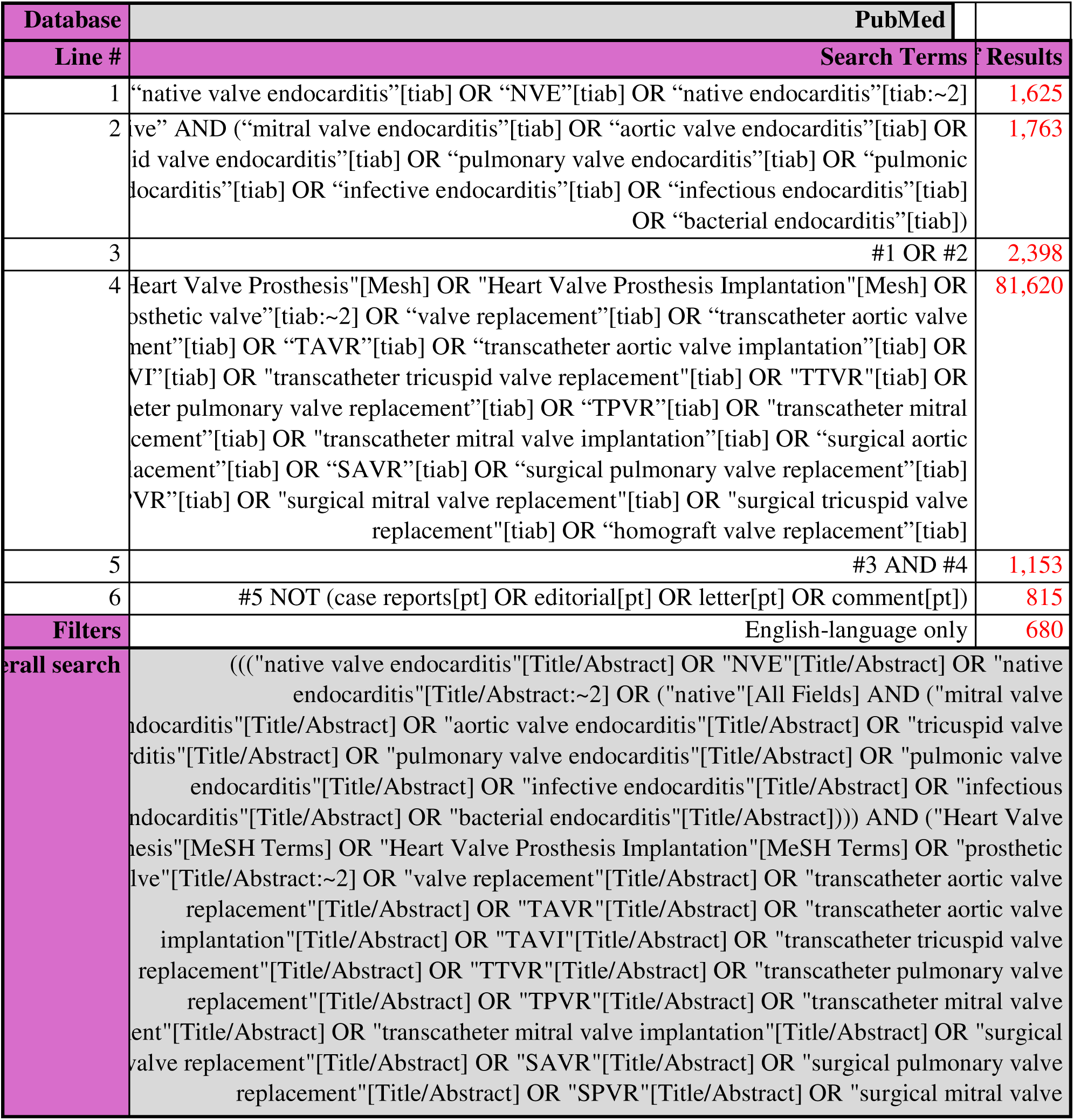
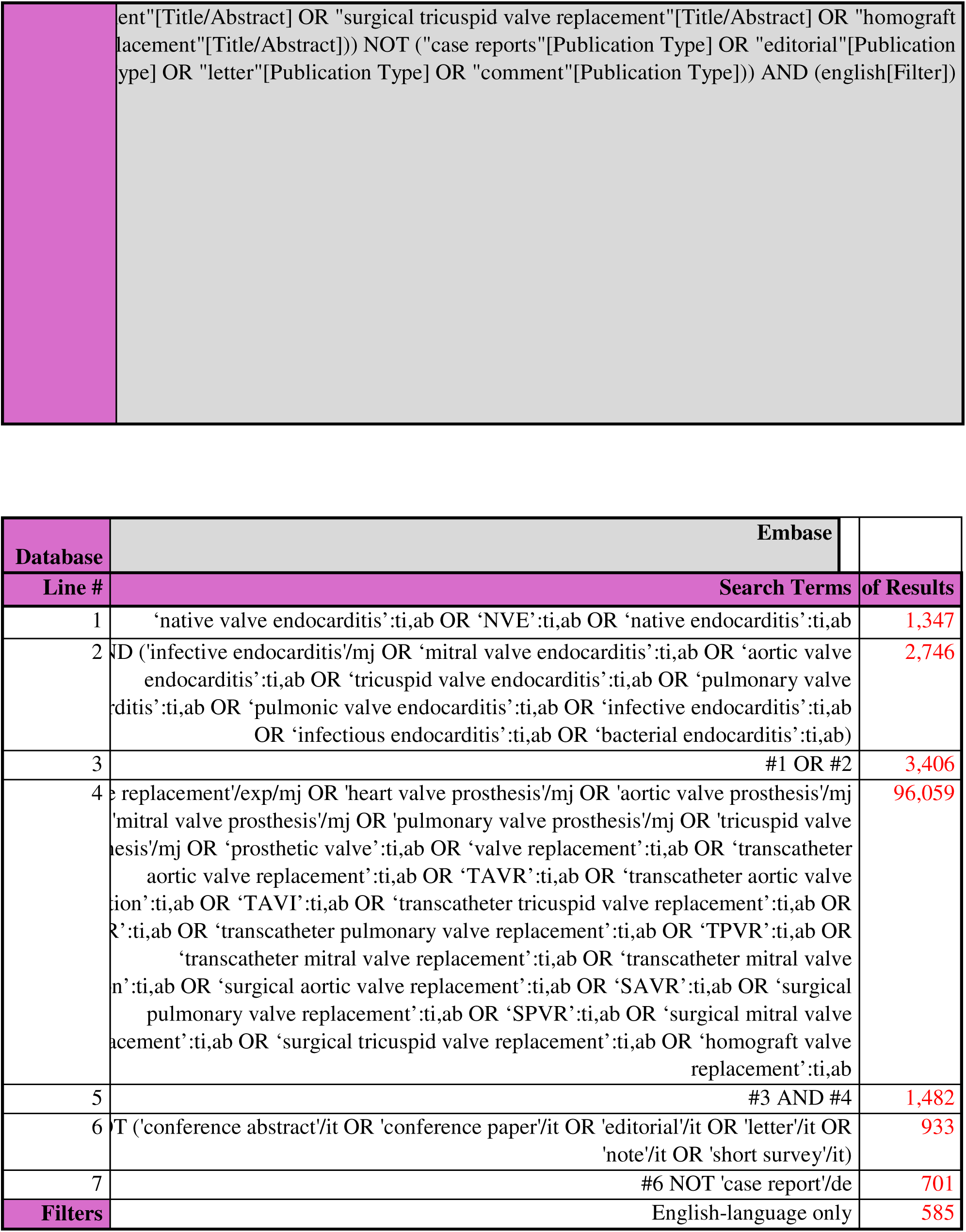
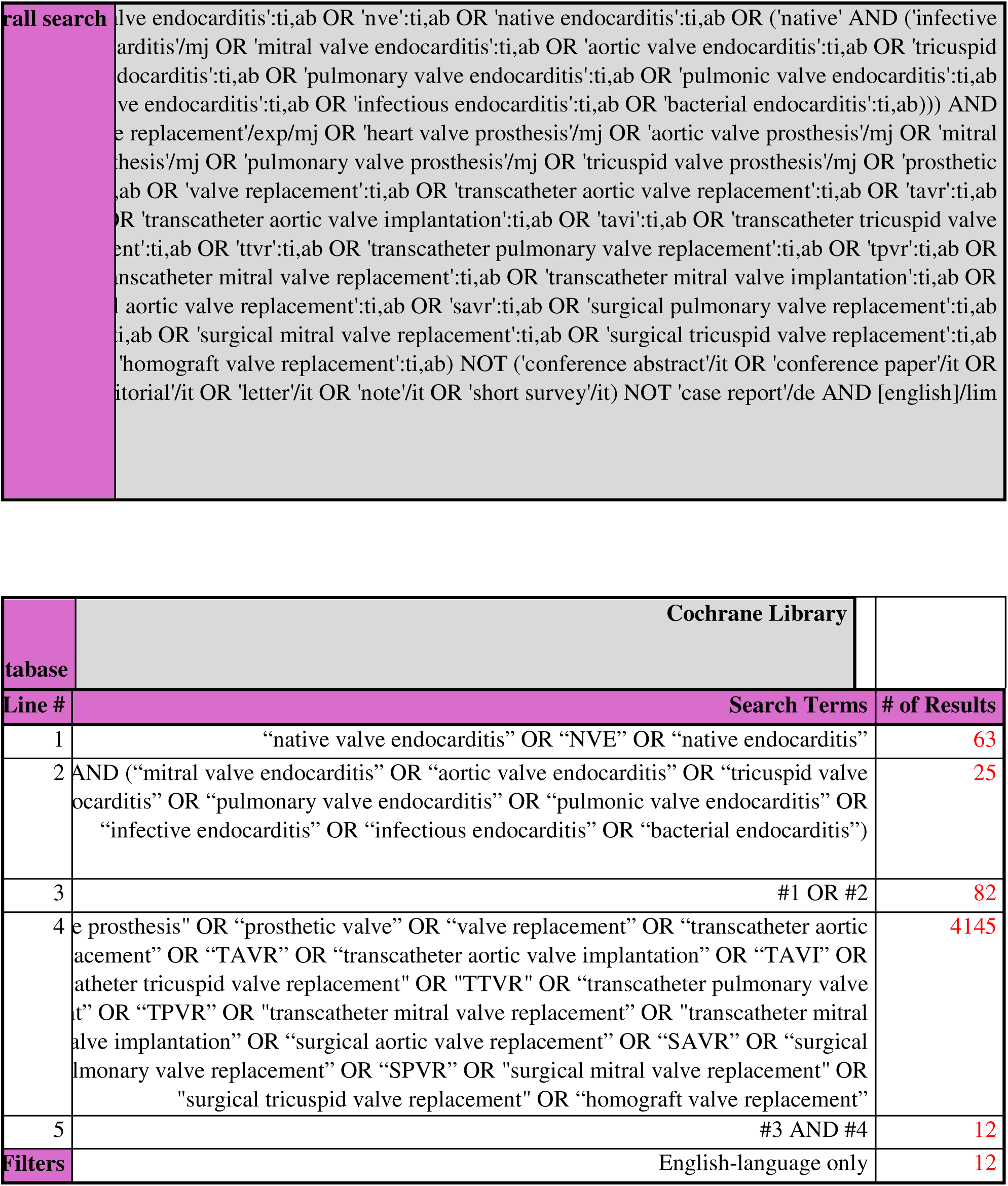
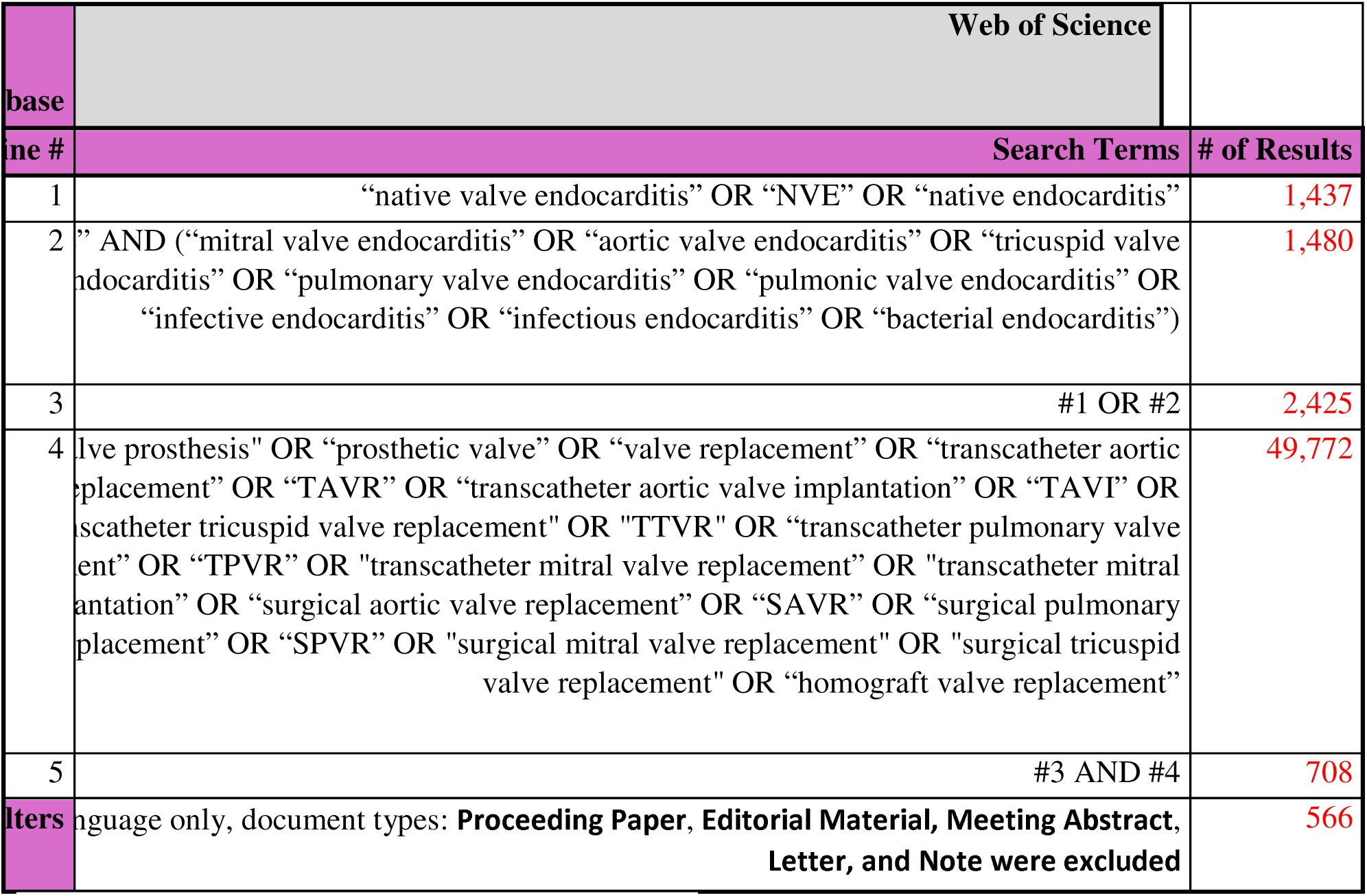
Search criteria in Pubmed, Embase, Cochrane, and Web of Science databases.

### Definition and Distinction of Valvular Endocarditis

The definition and diagnosis of IE adopted in our review were symptoms/signs of IE with evidence of organism growth in serial blood cultures and echocardiographic evidence of any of the following lesions in the endocardium - vegetation, abscess, aneurysm, fistula, pseudoaneurysm, or dehiscence. Distinction between PVE, NVE, and nonvalvular IE was made according to whether IE after PVI was reported on a prosthetic valve only versus prosthetic and native valve versus native valve only versus nonvalvular locations. This distinction was used to identify TAVIE (transcatheter aortic valve implant endocarditis), SAVIE (surgical aortic valve implant endocarditis), nonaortic prosthetic IE, combined PVE and NVE, INVE, and nonvalvular IE.

### Study selection

Figure 1 shows the PRISMA strategy for screening and exclusion using Covidence. All authors were aware of journal titles, authors, and sites. No limits were placed on study date or design except that conference abstracts, editorials, and commentaries were excluded. Case reports, case series, and meta analyses where included were done for reference purposes only and not for data analyses. Non-English studies were excluded. Review and cohort articles describing outcomes/results/events after PVI were included. After screening of abstracts, full texts were reviewed and, if meeting the criteria for INVE, were included for data extraction. References in these texts were also examined and included for data extraction if they met the criteria for INVE.

**Figure 1.**
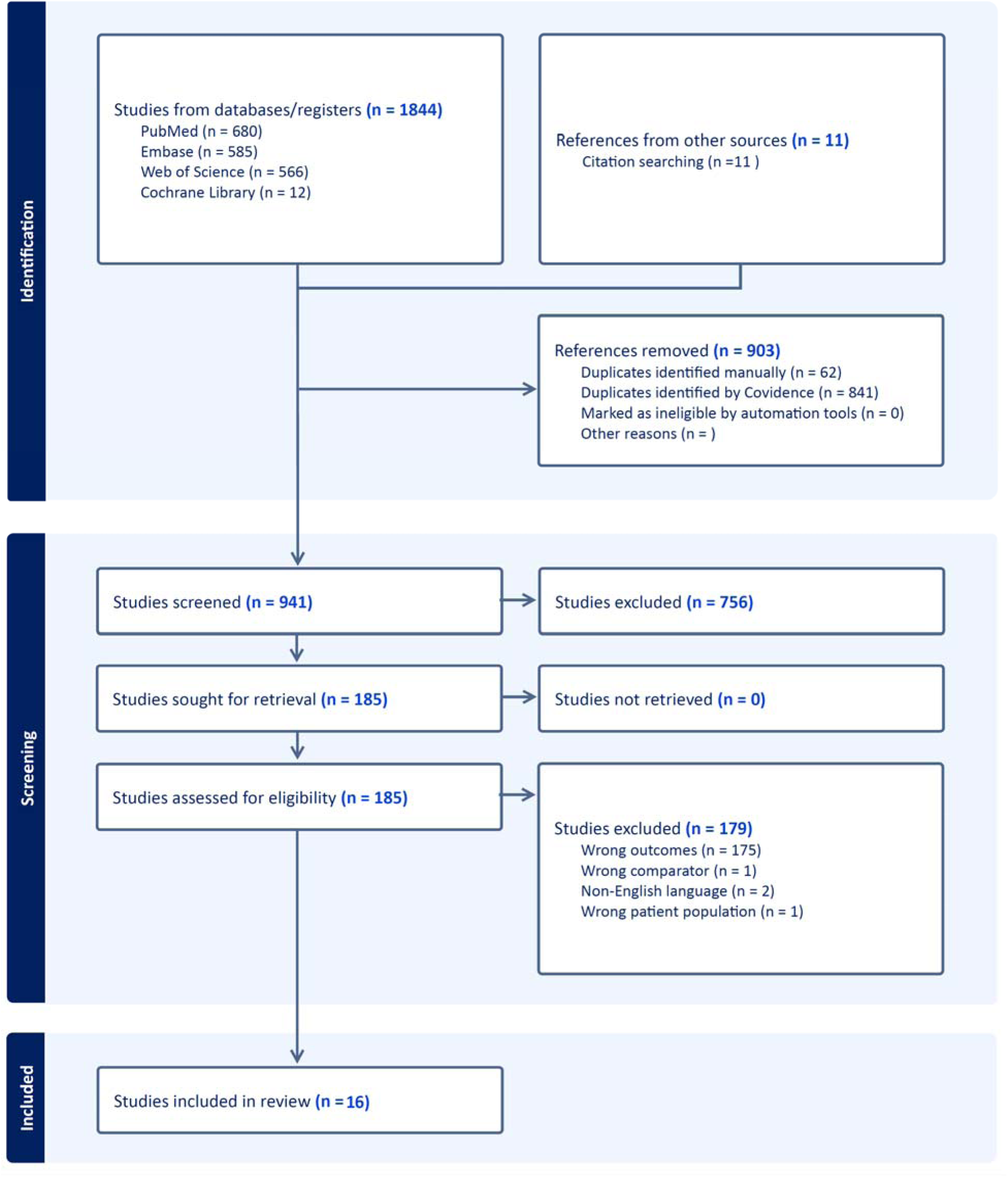
Prisma Search. Search results generated from Covidence using PRISMA protocol. Breakdown of results from four databases and criteria used to acquire final studies for review (n = 16)

### Data extraction

Articles in the final set were examined and data on several categories were identified. Specifically IE incidence after PVI, age groups affected, gender affected, comorbidities, PVI type, method of PVI, pathogens, left ventricular ejection fractions, treatment type, follow up periods, mortality rates, and causes of mortality were determined for PVE and INVE groups. Combined PVE and NVE cases were included in the PVE group. Nonvalvular IE cases were excluded. Data was acquired directly through articles, contact with the authors of the articles, and the IE after TAVR registry^5^. All data are available in the tables with the manuscript. All authors contributed to data acquisition.

## Results

### Search outcomes

The search yielded 952 studies after removal of duplicates in Covidence. In these 952 studies, abstracts that described the outcomes of PVI were identified and moved into full text review. Out of 196 full texts on PVI outcomes, 16 matched the criteria for INVE. These studies were manually reviewed for INVE populations from the same registries. References in these studies were also reviewed. Ultimately, INVE cases reporting non TAVIs and non SAVIs were excluded from data analyses since very few cases were identified. INVE cases from 8 studies on TAVI and SAVI were included in the final analysis (Table 2). These studies comprised of retrospective reviews and complete prospective cohorts. A p value less than 0.05 was considered significant.

**Table 2.**
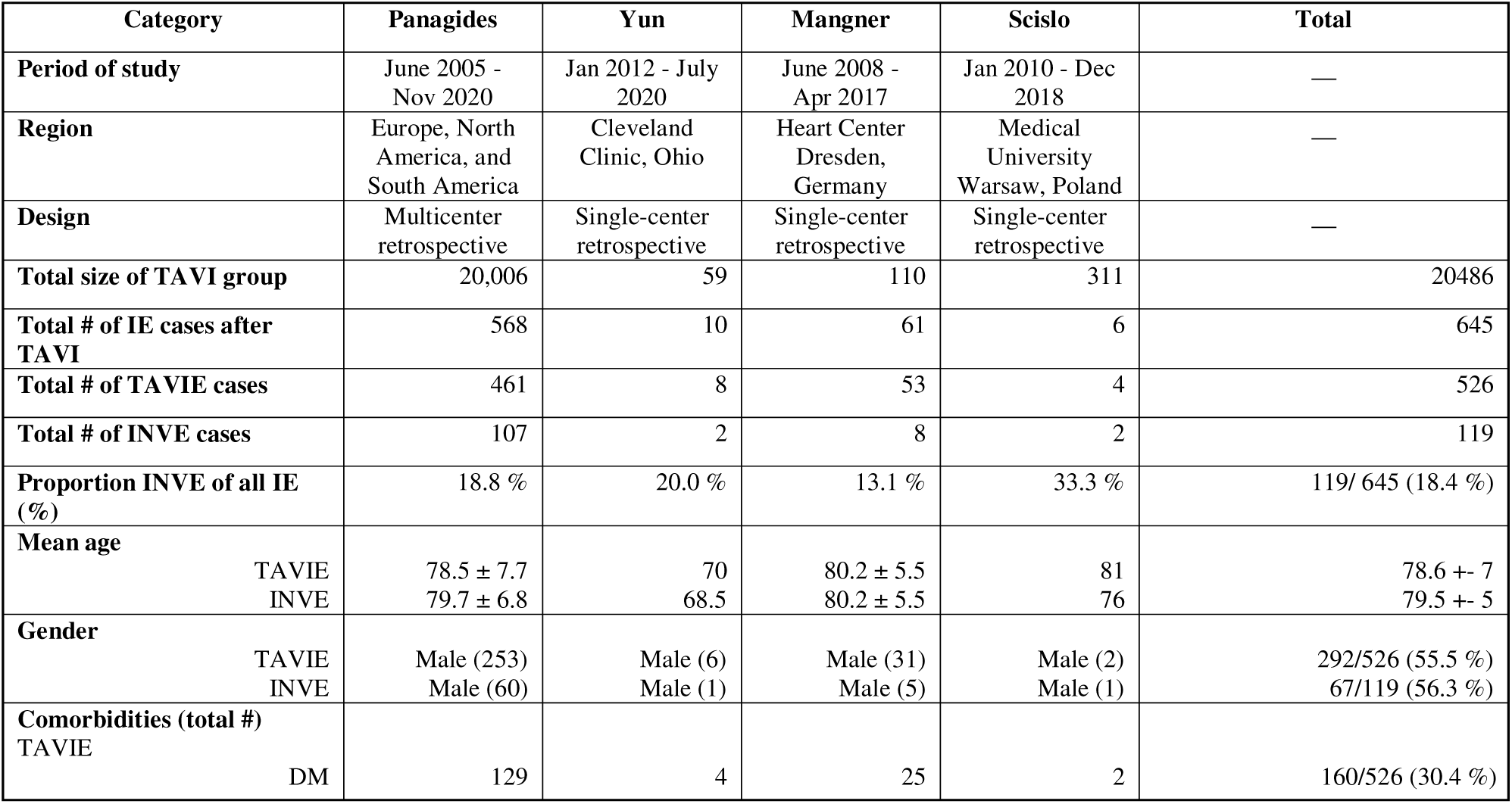
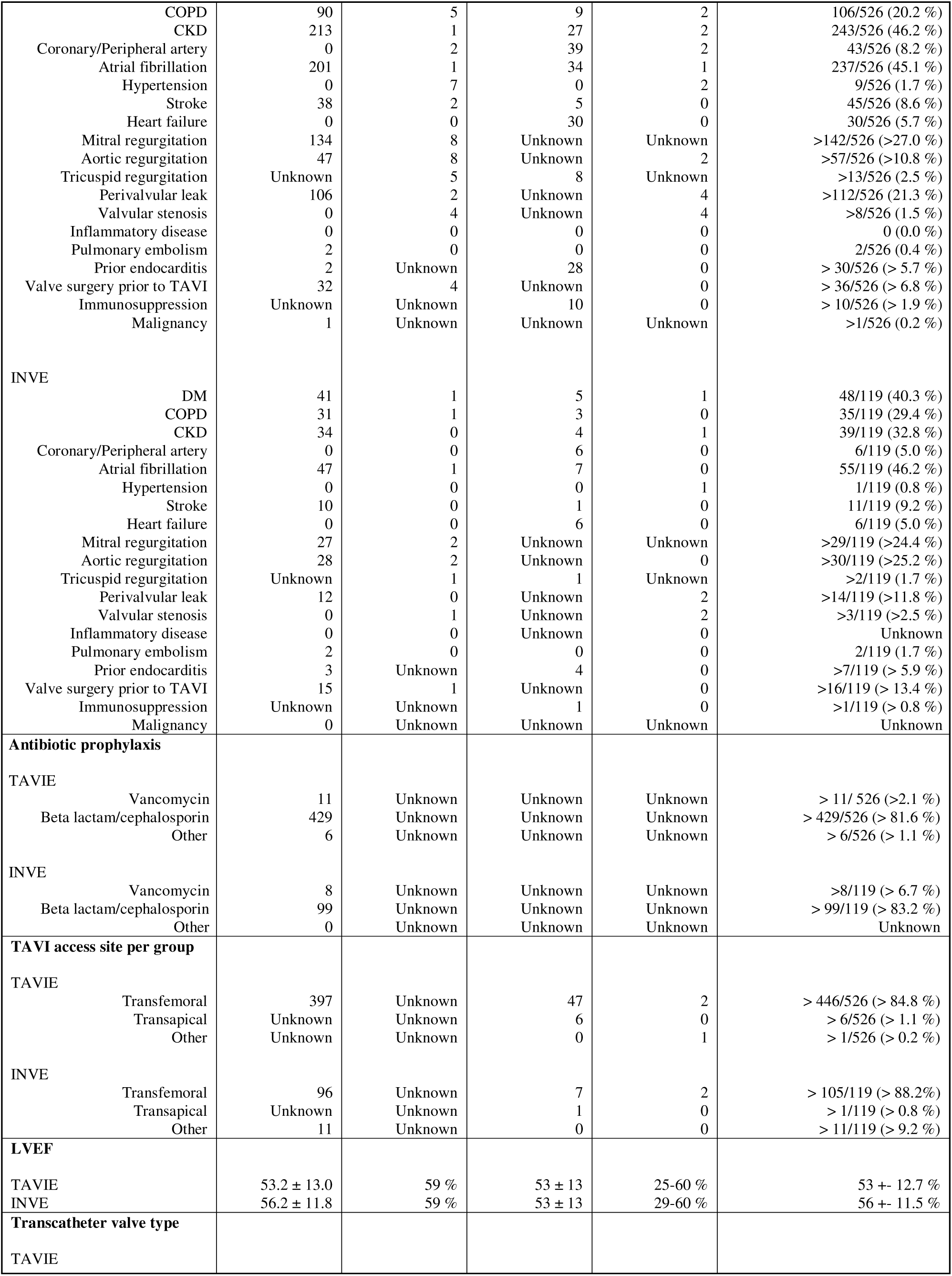
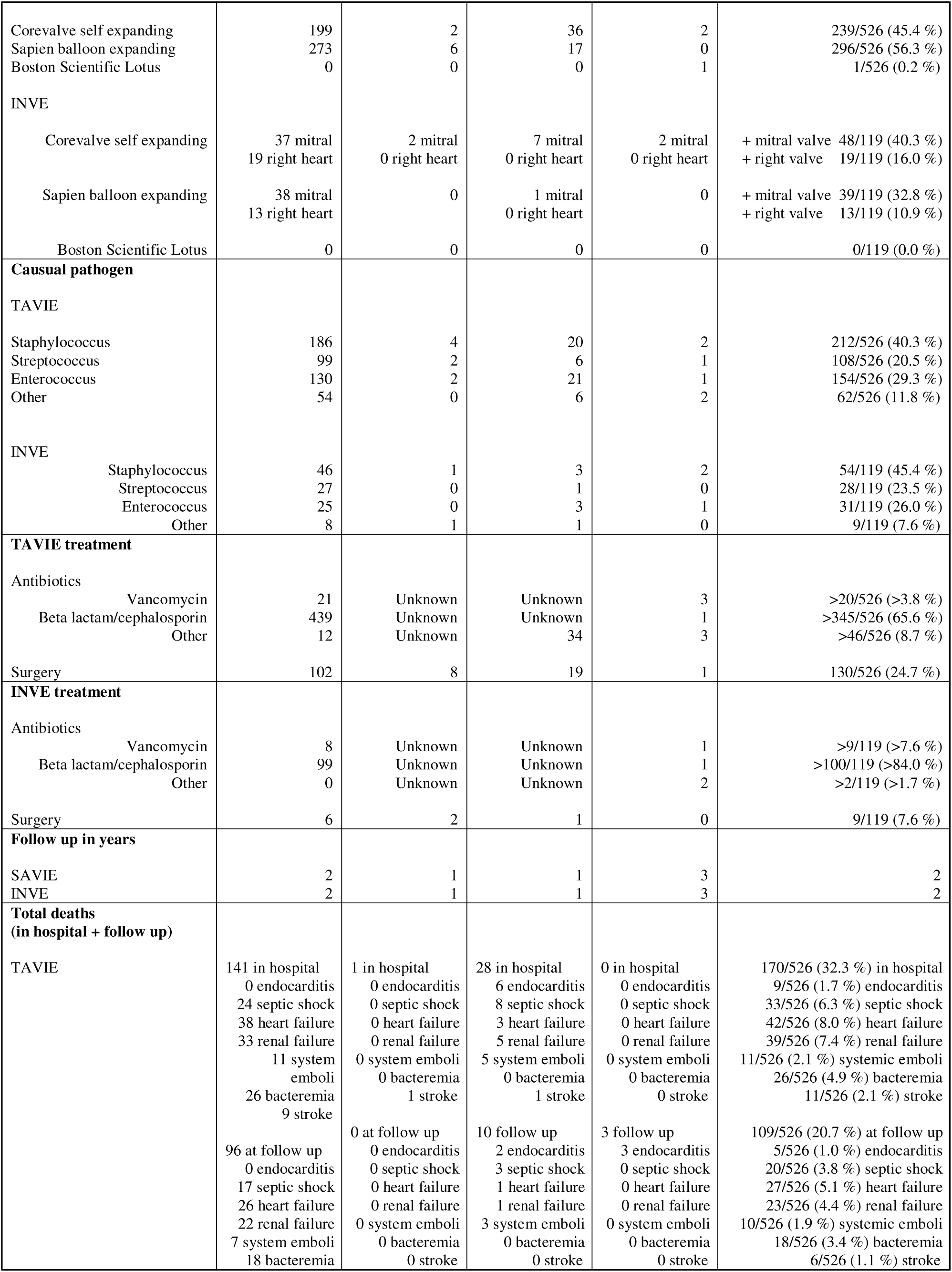
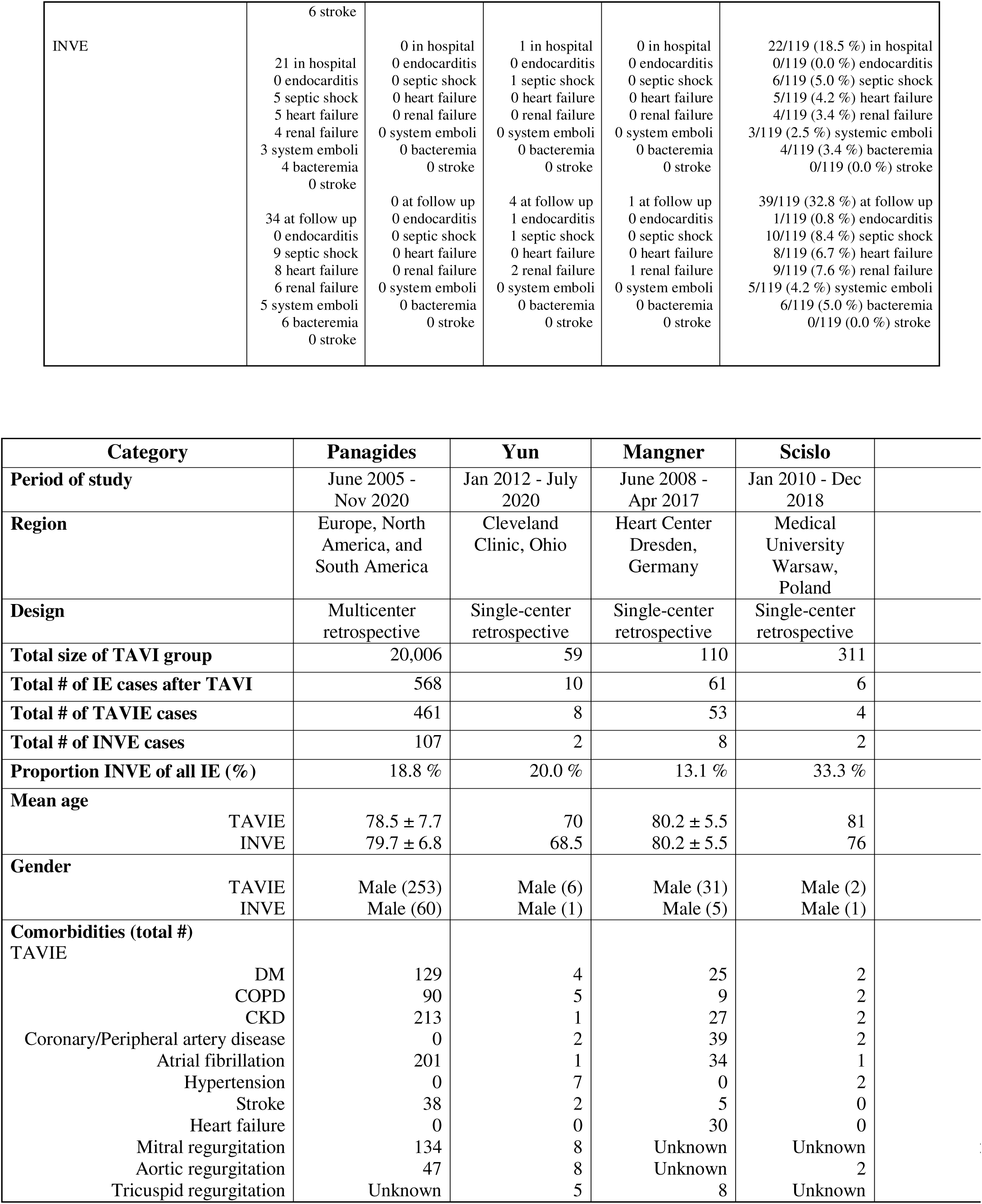
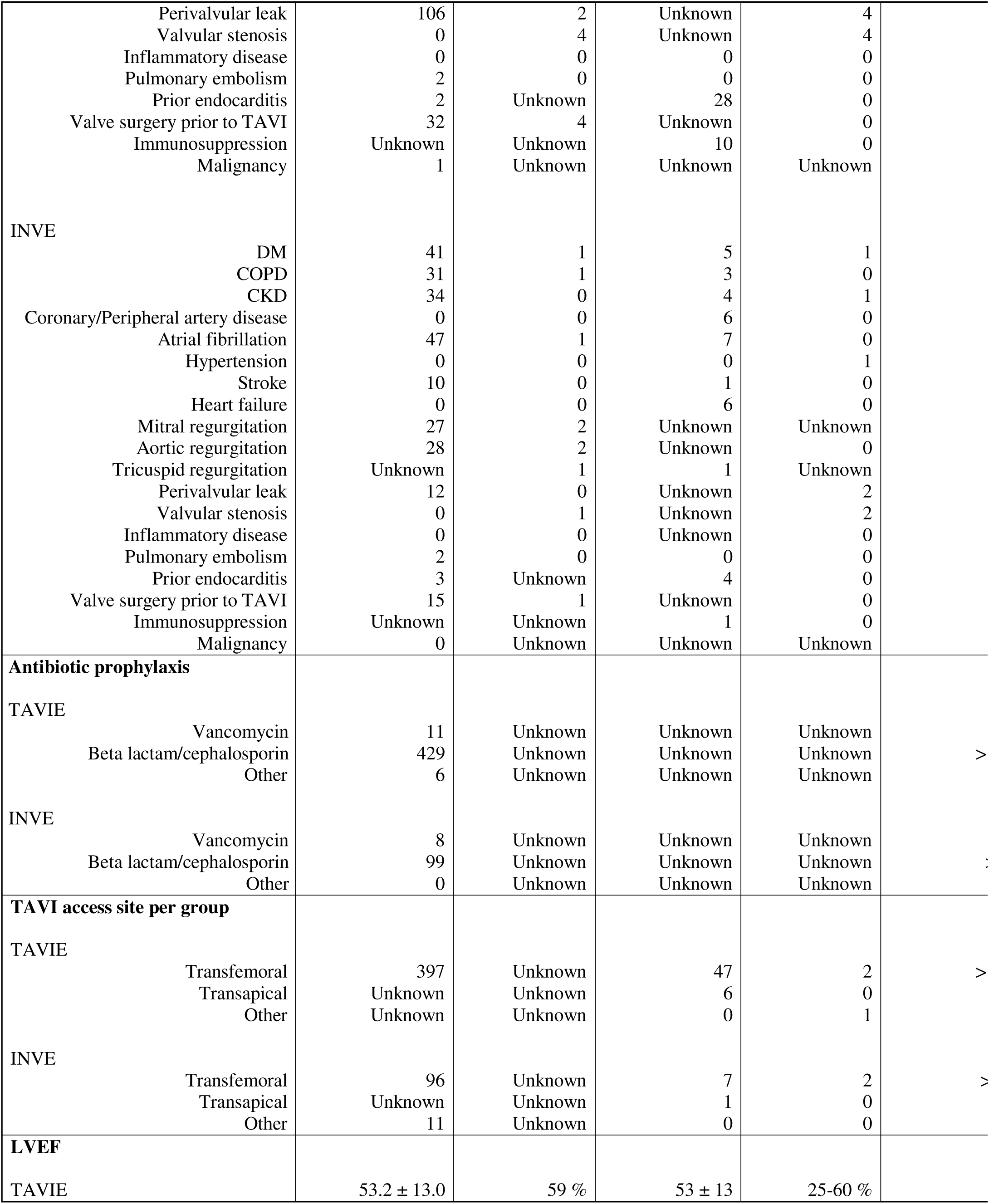
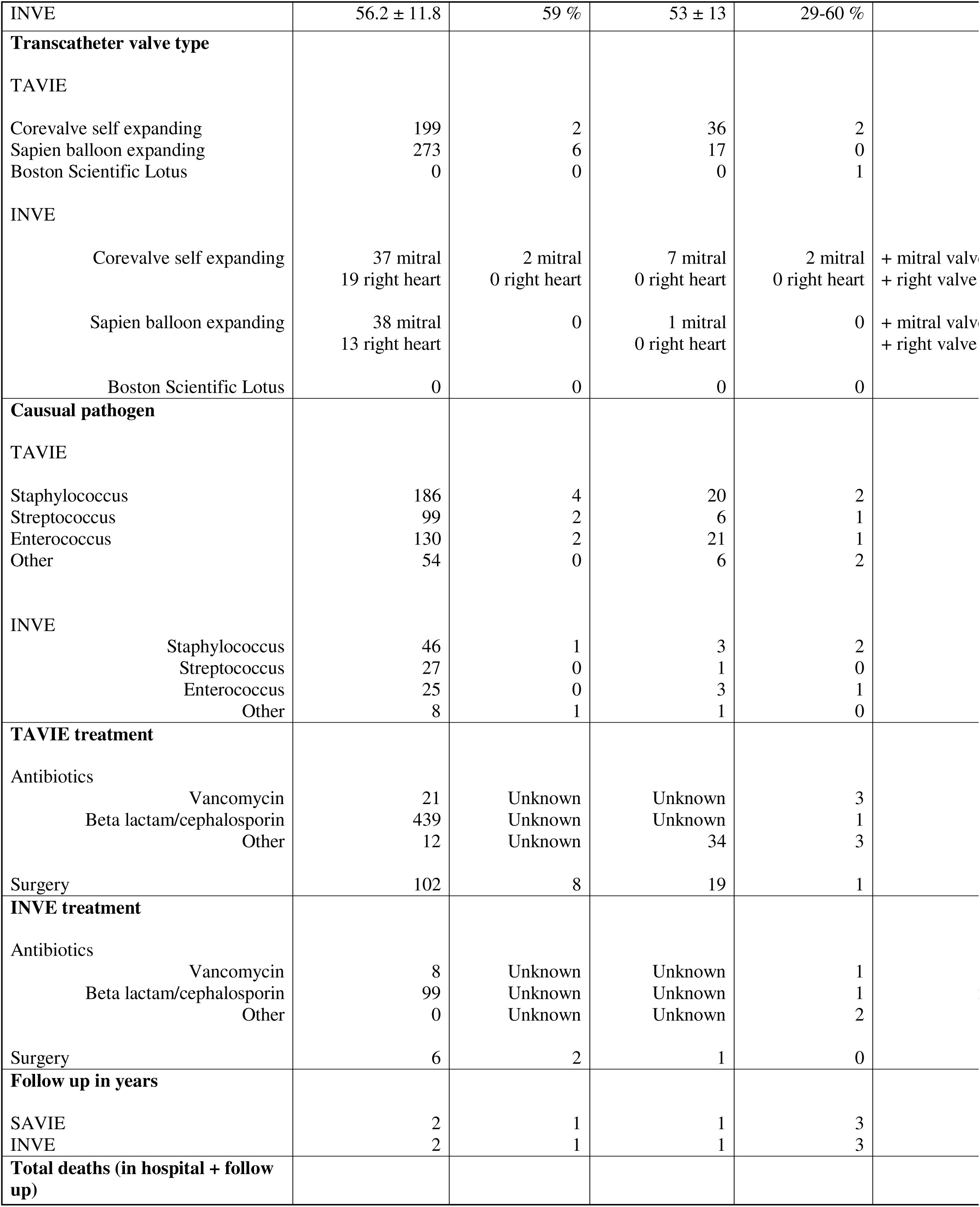
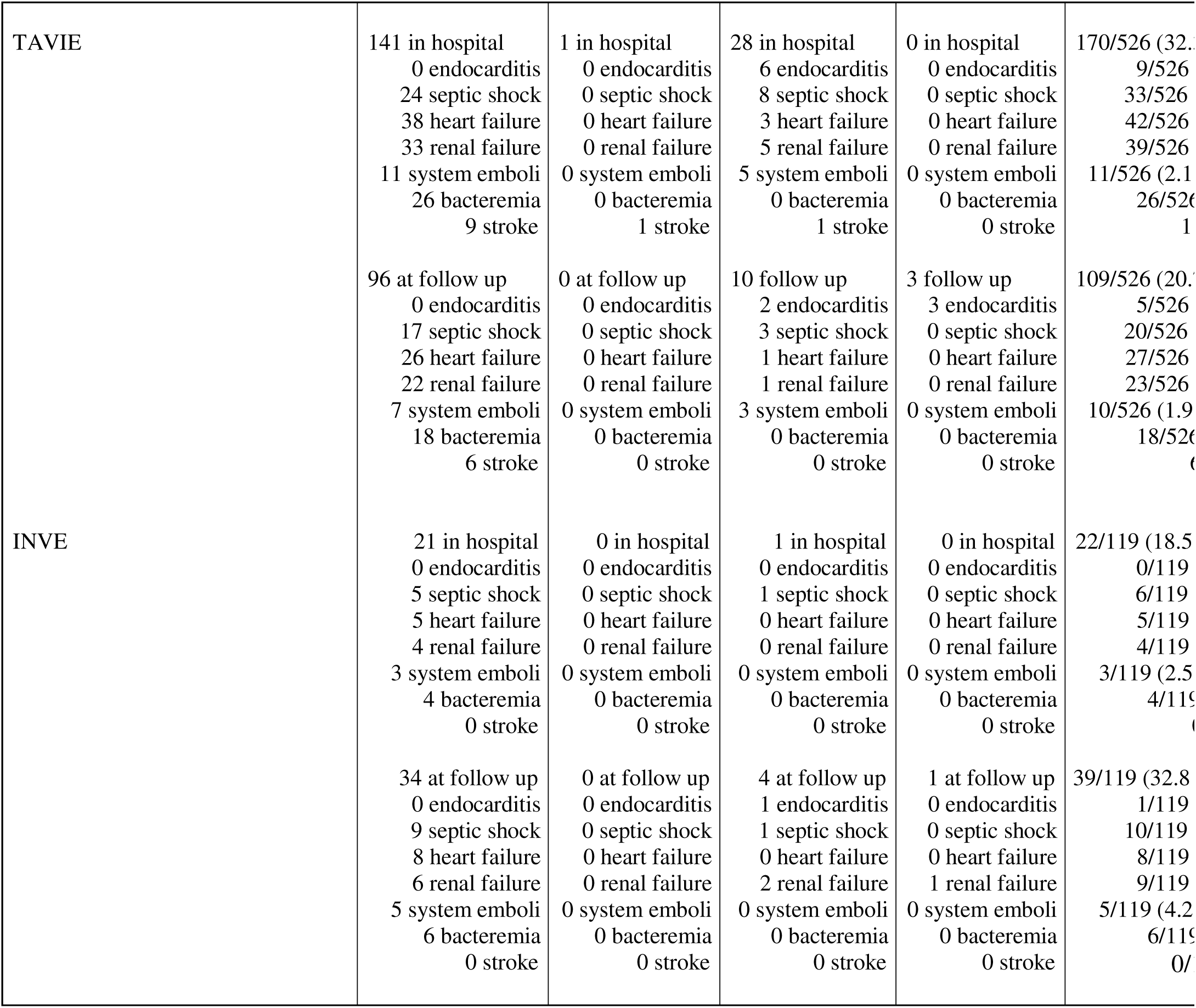
Summary of findings in TAVI group. Summary of categorical findings for TAVIE AND INVE in TAVI studies (n = 4). Missing data is marked as “unknown”

### Risk of bias

All 8 articles were of good quality based on assessment of selection, comparability, and exposure categories in the Newcastle Ottawa assessment. However, some data on the categories of interest to our study were not reported; these data were marked as “unknown” or “?”in Tables 2 and 4. The INVE group was represented in all studies. The studies obtained patient and outcome information from institutional medical records. All studies assessed outcomes and ensured adequate follow-up. Follow-up time was sufficient for outcomes to occur in all cohorts. Notably, one study drew data from an international registry consisting of multicenter databases while remaining studies drew data from unicenter databases. The discrepancies in population selection and size raise the possibility that associations close to cutoff for significance (p < 0.05) may be stronger or weaker if comparable populations are analyzed. Moreover, associations that are insignificant (p > 0.05) may also be different if similar populations are analyzed.

### INVE after TAVI

INVE was observed in 119/645 (18.4 %) cases of valvular IE after TAVI. The mean age for the INVE group was 79.5 +- 5 years and comparable to the TAVIE group (Table 2). In both INVE and TAVIE, cases were more frequent in males at 56.3 % and 55.5 % respectively. Comorbidity rates in INVE and TAVIE were similar except that diabetes (DM) was present in 40.3 % INVE vs 30.4 % TAVIE (OR 1.55, CI 1.03 - 2.33, p = 0.037). Chronic obstructive pulmonary disease (COPD) was present in 29.4 % INVE vs 20.2 % TAVIE (OR 1.65, CI 1.05 - 2.58, p = 0.027). Chronic kidney disease (CKD) was present in 32.8 % INVE vs 46.2 % TAVIE (OR 0.57, CI 0.37 - 0.86, p = 0.008).

Transfemoral access was utilized for TAVI prior to most cases of INVE and TAVIE with an insignificantly higher rate in the INVE group. Beta lactam antibiotic prophylaxis was carried out in at least 80% of both groups with other antibiotics contributing the remaining 20%. No concurrent infections were noted in either group. Moreover, there were few instances of prior endocarditis or prior valvular procedures in either group and where present occurred at comparable rates. Mean left ventricular ejection fraction (LVEF) in INVE was 56 +-11.5 %, similar to TAVIE. Corevalves were more frequently associated with INVE than TAVIE at 56.3 % vs 45.4 % (OR = 1.55, CI 1.04 - 2.31, p = 0.032). The valvular distributions of INVE were 87/119 (73.1%) on mitral valves and 32/119 (26.9%) on right heart valves. The specific right heart valve locations of the latter were not identifiable. Infections in INVE and TAVIE were from gram positive flora such as staphylococci, streptococci, and enterococci at comparable rates. The sources of these pathogens were unknown in most of the cases, although gastrointestinal and urinary tract were considered likely points. INVE cases required surgery less frequently than did TAVIE cases at 7.6 % vs 24.7 % (RR = 0.31, CI 0.16 - 0.58, p < 0.001).

Ultimately, TAVIE and INVE cases were followed for approximately 2 years. INVE mortalities were lower in hospital at 21.0 % INVE vs 32.3 % TAVIE (RR 0.65, CI 0.45 - 0.94, p = 0.016). INVE mortalities were higher at follow up at 30.3 % INVE vs 20.7 % TAVIE (RR 1.47, CI 1.07 - 2.03, p = 0.021). Mortality rates due to any particular cause were similar across both groups in hospital and at follow up (Table 3).

**Table 3.**
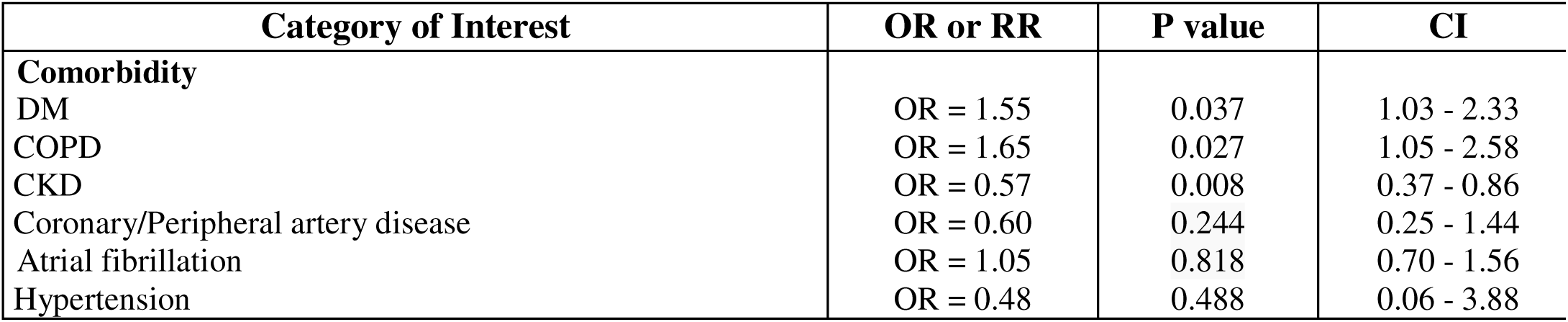
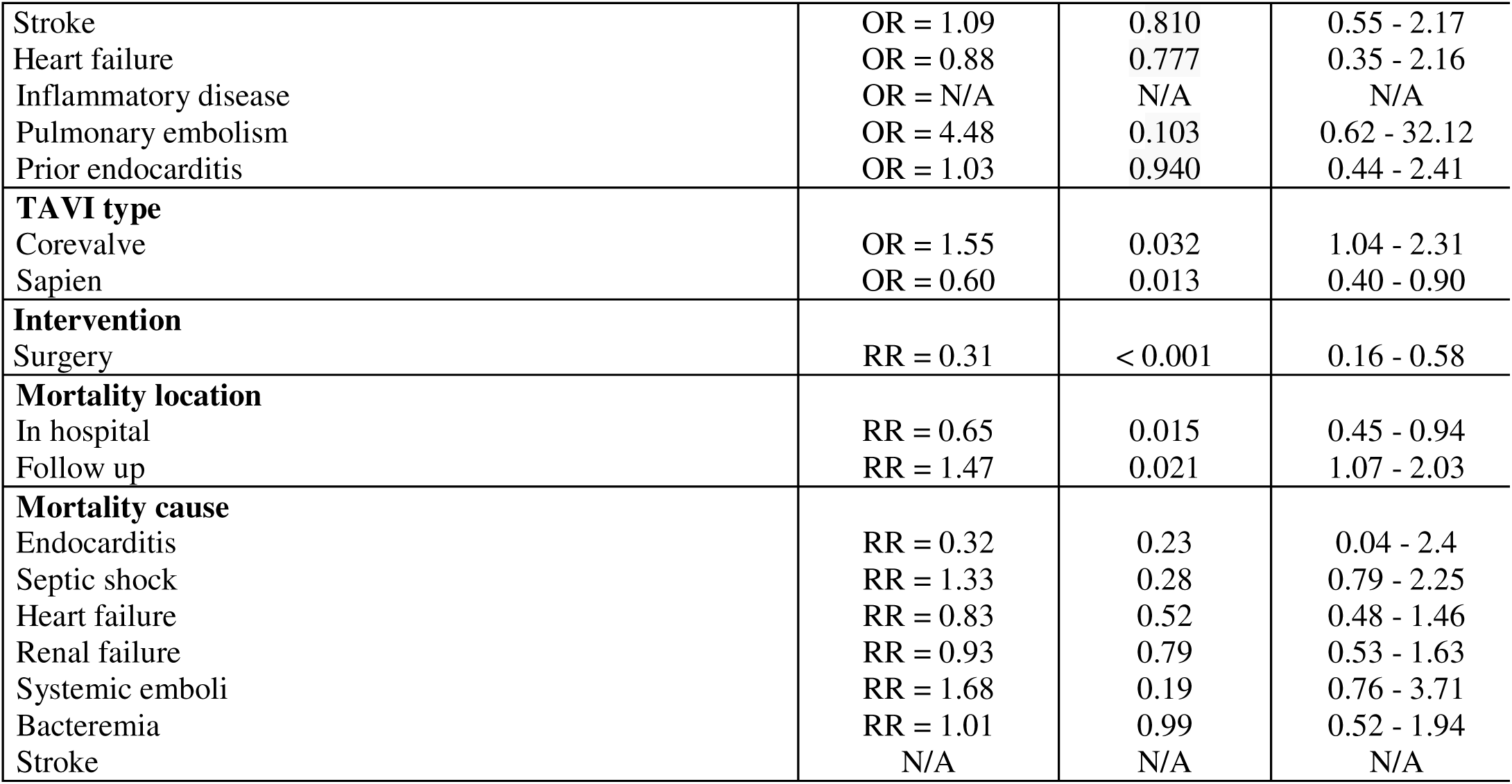
Statistical comparison of categories between TAVIE and TAVIE INVE groups. Categories with significant vs non-significant differences between INVE and TAVIE after chi-square analyses with odds ratio (OR), relative risk (RR), P value, and confidence interval (CI).

| Category of Interest | OR or RR | P value | CI |
| --- | --- | --- | --- |
| **Comorbidity** |  |  |  |
| DM | OR = 1.55 | 0.037 | 1.03 - 2.33 |
| COPD | OR = 1.65 | 0.027 | 1.05 - 2.58 |
| CKD | OR = 0.57 | 0.008 | 0.37 - 0.86 |
| Coronary/Peripheral artery disease | OR = 0.60 | 0.244 | 0.25 - 1.44 |
| Atrial fibrillation | OR = 1.05 | 0.818 | 0.70 - 1.56 |
| Hypertension | OR = 0.48 | 0.488 | 0.06 - 3.88 |
| Stroke | OR = 1.09 | 0.810 | 0.55 - 2.17 |
| Heart failure | OR = 0.88 | 0.777 | 0.35 - 2.16 |
| Inflammatory disease | OR = N/A | N/A | N/A |
| Pulmonary embolism | OR = 4.48 | 0.103 | 0.62 - 32.12 |
| Prior endocarditis | OR = 1.03 | 0.940 | 0.44 - 2.41 |
| **TAVI type** |  |  |  |
| Corevalve | OR = 1.55 | 0.032 | 1.04 - 2.31 |
| Sapien | OR = 0.60 | 0.013 | 0.40 - 0.90 |
| **Intervention** |  |  |  |
| Surgery | RR = 0.31 | < 0.001 | 0.16 - 0.58 |
| **Mortality location** |  |  |  |
| In hospital | RR = 0.65 | 0.015 | 0.45 - 0.94 |
| Follow up | RR = 1.47 | 0.021 | 1.07 - 2.03 |
| **Mortality cause** |  |  |  |
| Endocarditis | RR = 0.32 | 0.23 | 0.04 - 2.4 |
| Septic shock | RR = 1.33 | 0.28 | 0.79 - 2.25 |
| Heart failure | RR = 0.83 | 0.52 | 0.48 - 1.46 |
| Renal failure | RR = 0.93 | 0.79 | 0.53 - 1.63 |
| Systemic emboli | RR = 1.68 | 0.19 | 0.76 - 3.71 |
| Bacteremia | RR = 1.01 | 0.99 | 0.52 - 1.94 |
| Stroke | N/A | N/A | N/A |

### INVE after SAVI

INVE was observed in 9/138 (6.5 %) cases of valvular IE after SAVI. The mean age for the SAVIE group was 59.9 ± 15.8 years and for INVE group was 64.4 ± 3.8 years (Table 4). Cases were reported more frequently in males than females. Comorbidities observed with both SAVIE and INVE included diabetes, hypertension, heart failure, mitral regurgitation, aortic regurgitation, prior endocarditis, prior valve surgery, and malignancy. In the SAVIE group, staphylococcus was source of infection in 44% of cases and surgical management was required in about one third of the cases. Mortality rates in the SAVIE group were 18 % in hospital and 7 % at follow up due to endocarditis complications. Data on the INVE group was limited. It should be noted however that in 2 of the 9 INVE cases, one was a tricuspid INVE associated with rheumatic disease and the source of infection was the uninfected SAVI. The other case was a mitral INVE with infection suspected from cardiac catheterization and uninfected SAVI. A formal statistical analysis of the categories of interest in the SAVI INVE and SAVIE groups was not possible given the small sizes and incomplete data on the groups.

**Table 4.**
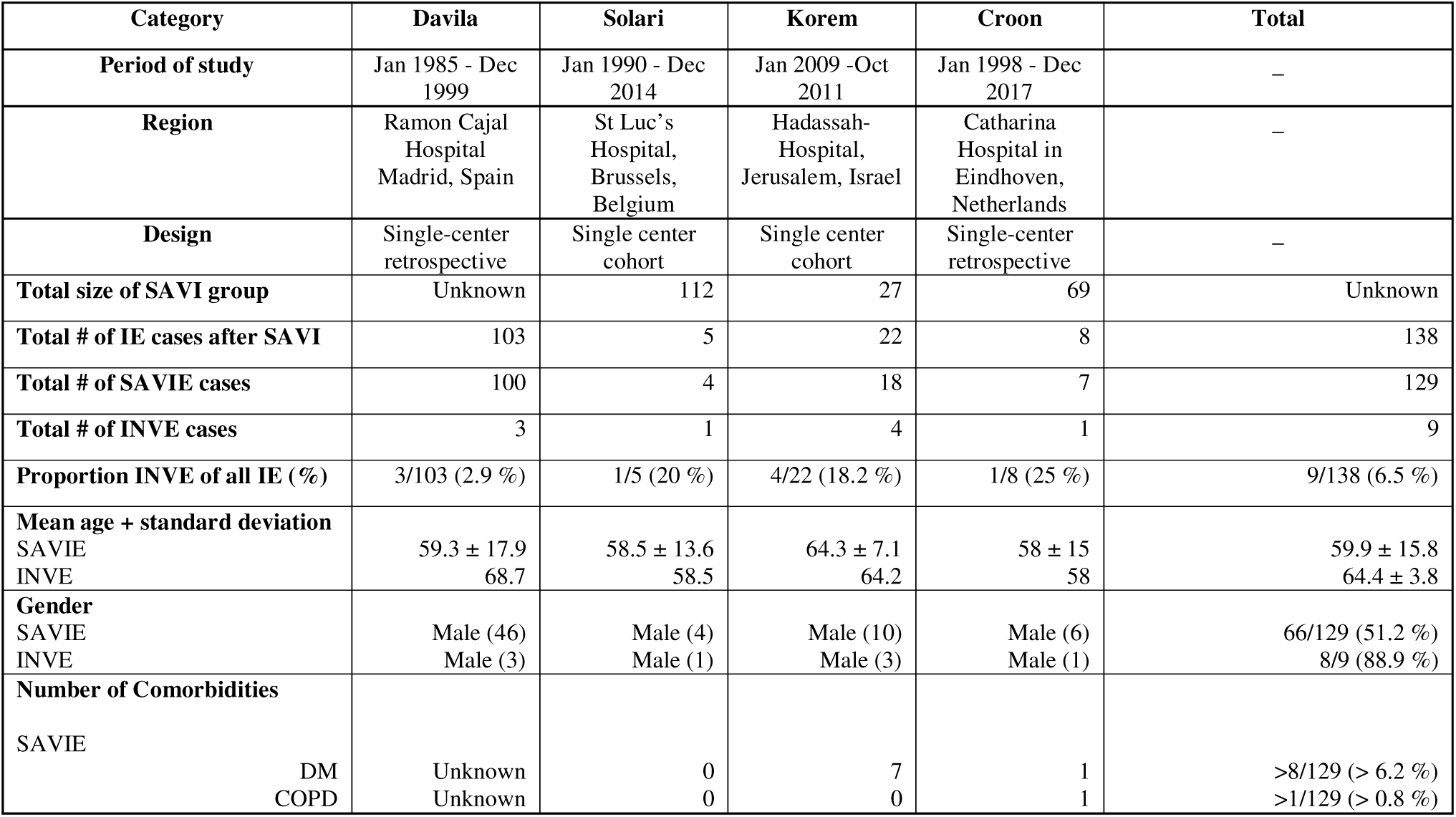
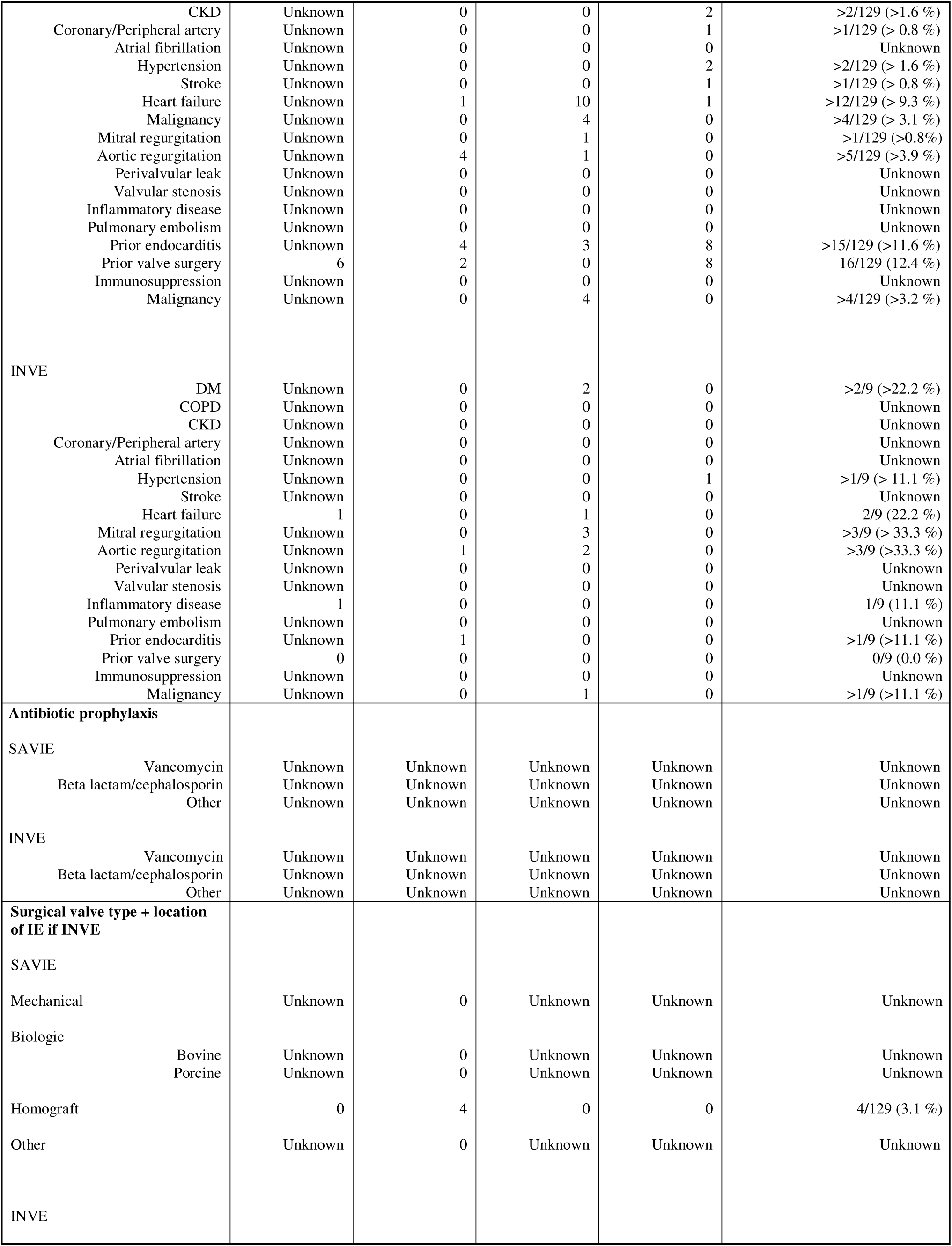
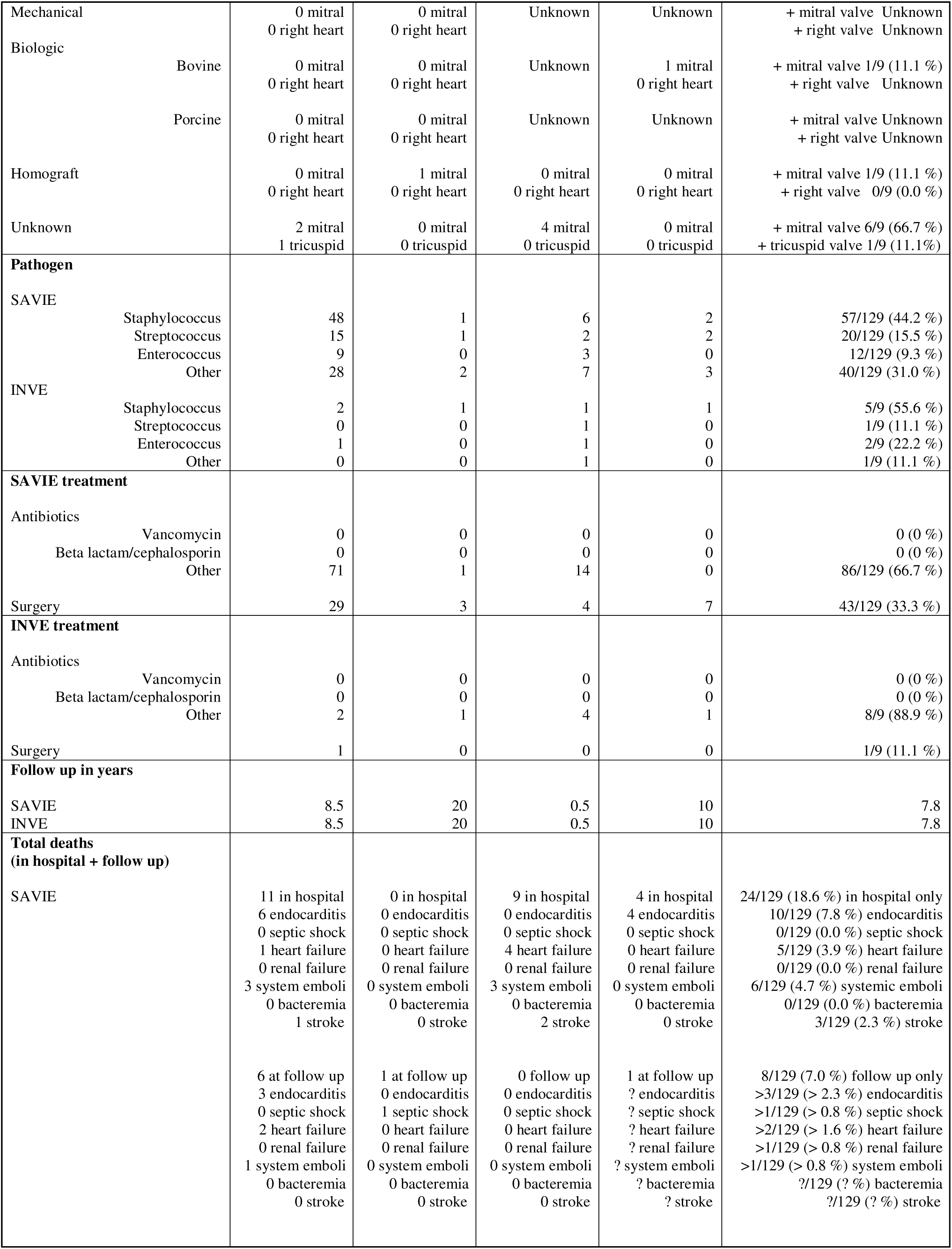
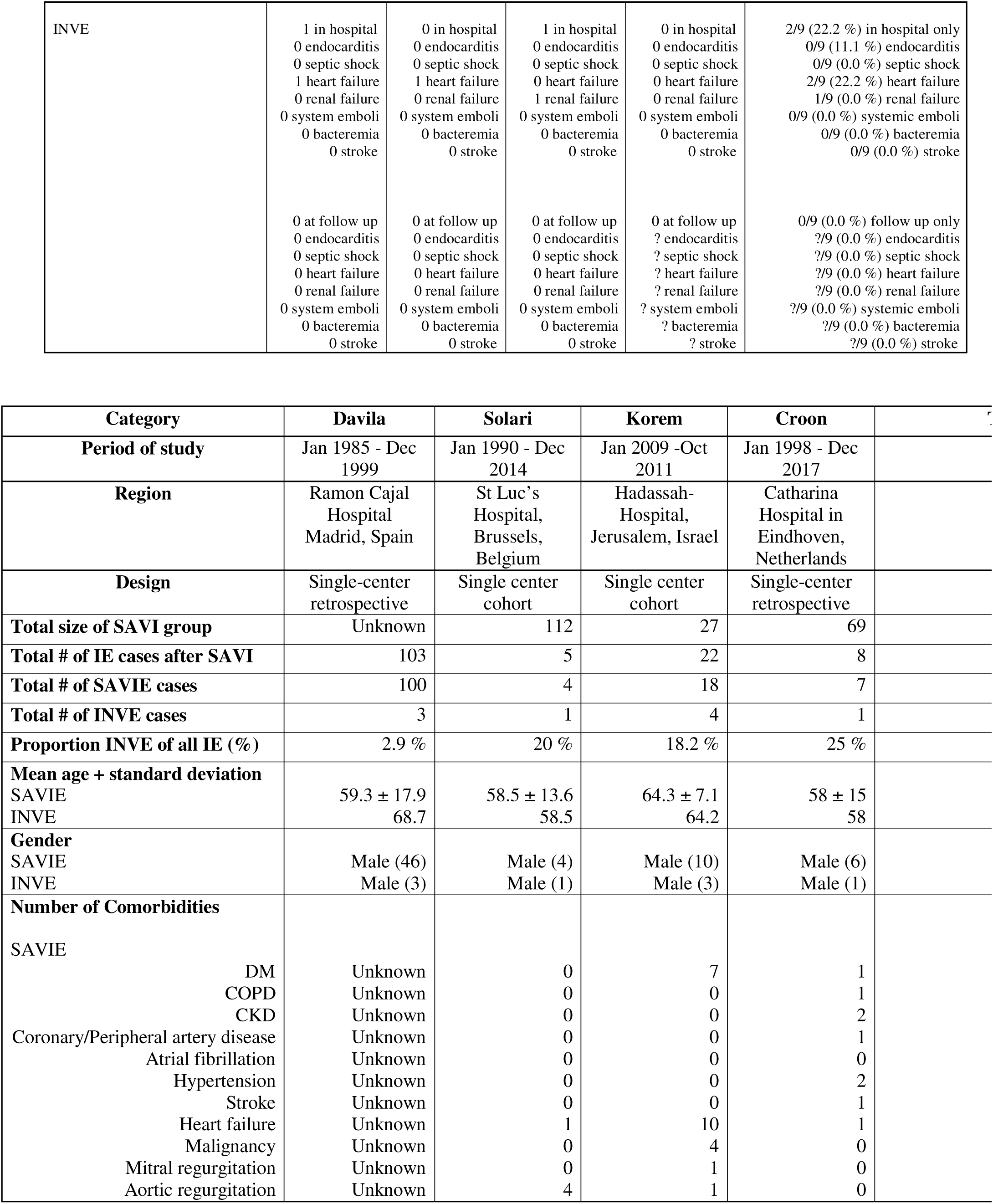
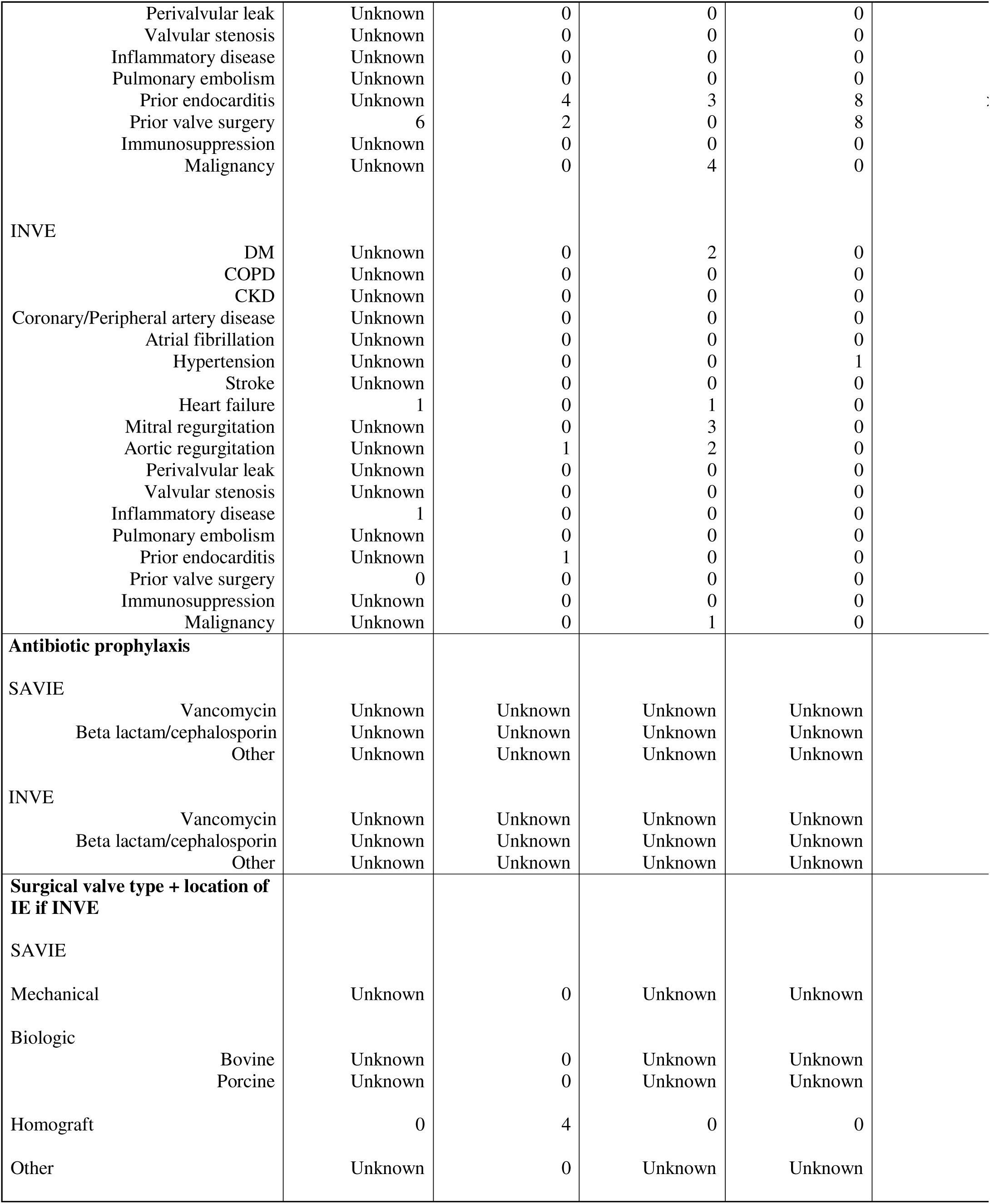
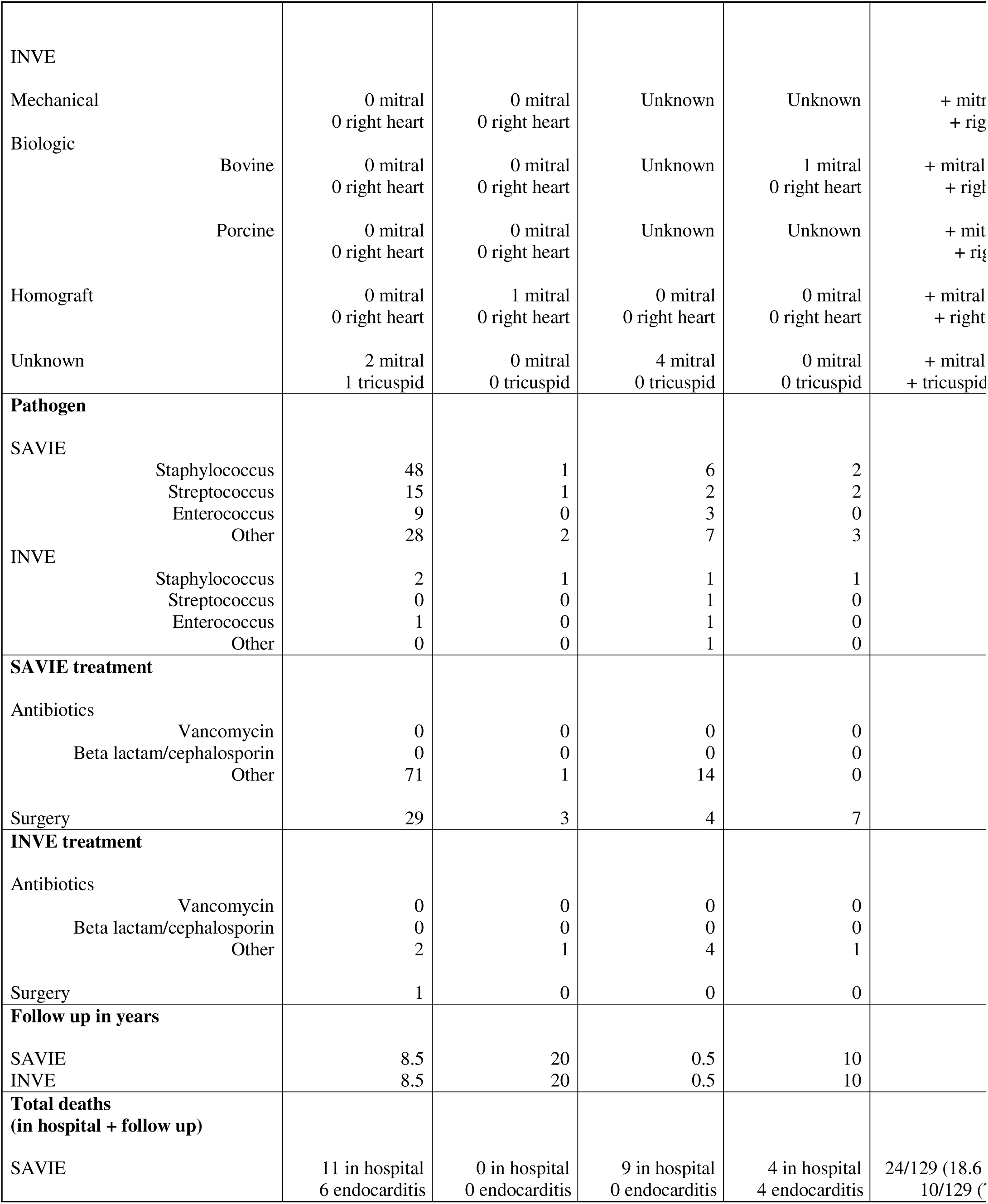
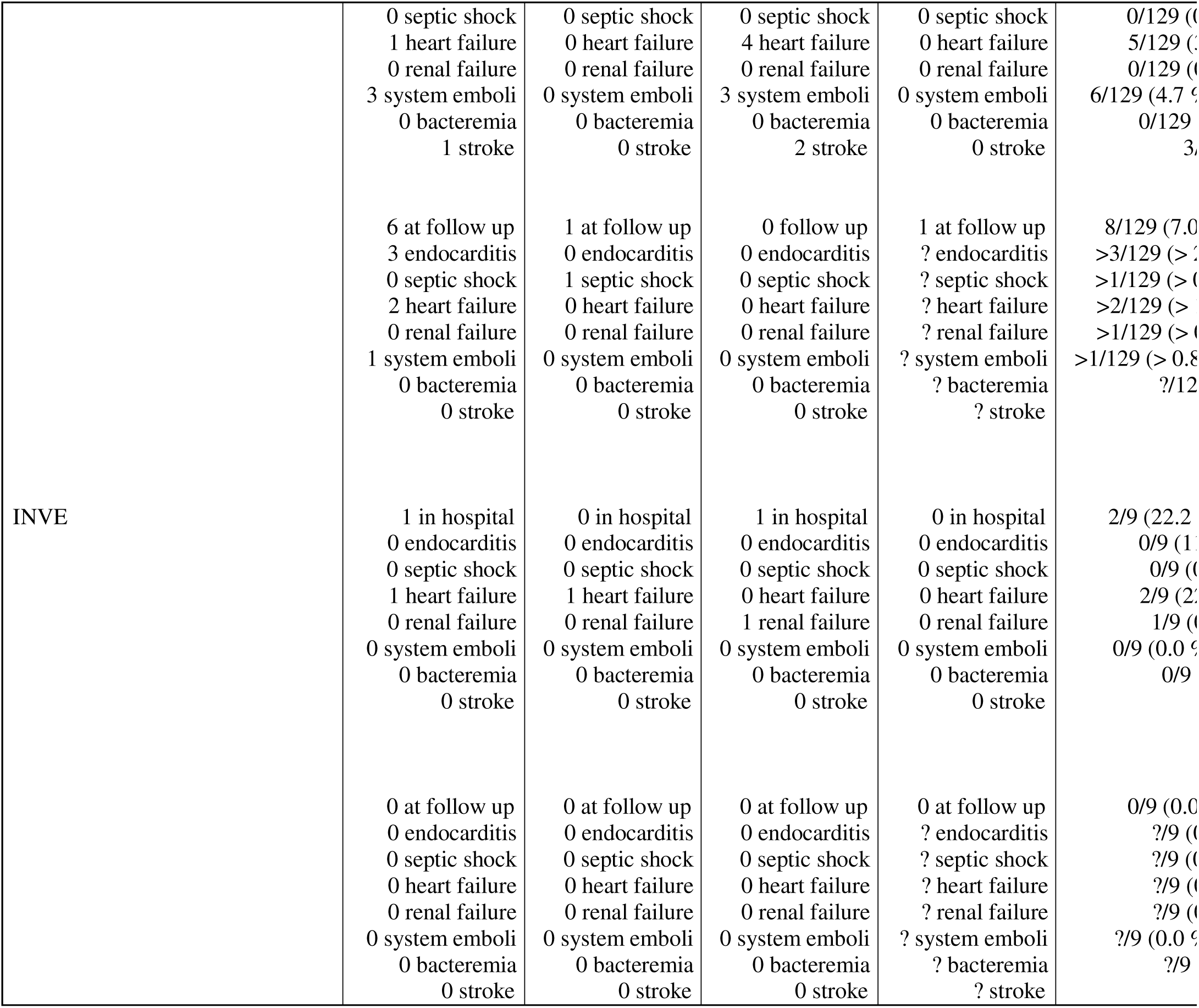
Summary of findings in SAVI groups. Summary of categorical findings for SAVIE populations per final study (n = 4). Missing data is marked as “unknown” or “?”

One instance of mitral INVE after SAVI that was not included in the data analysis was a case report of a Brucella infection^19^. The patient was 58 year old male with history of severe aortic stenosis requiring SAVI. Details regarding antibiotic prophylaxis, surgical approach, prosthesis type, ejection fraction, and suspected cause of INVE were not provided. The patient was treated with antibiotic therapy alone without subsequent INVE complications and was alive at 1 year follow up.

### INVE in non aortic prostheses

A few studies in our search reported INVE subsequent to non-aortic prostheses. One study described a single case of mitral INVE subsequent to transcatheter pulmonic valve replacement^8^. In this retrospective single center study, the patient was a 54 year old female with history of end stage renal disease on dialysis, pulmonic stenosis, and pulmonic endocarditis requiring Sapien 3 prosthetic implantation. The prosthetic valve was free of infection and the dialysis line was determined to be the source for mitral INVE. The pathogen was methicillin sensitive staph aureus treated with antibiotics and surgical mitral replacement. The prognosis was good and patient was alive after 2 years of follow up.

Two studies reported on aortic INVE subsequent to mitral PVI. One study was a cohort following patients after mitral surgical PVI^12^. A total of 9 patients were identified with 8 developing endocarditis on the mitral PVI and 1 developing aortic INVE. The details on this aortic INVE patient including the patient’s age, gender, comorbidities, surgical approach of PVI, PVI type, source of INVE, infecting pathogen, and complications after treatment of INVE could not be identified. However, it was known that the patient was treated with antibiotic therapy alone without surgery. Moreover, the patient was followed for 10 years with INVE developing 8 years after PVI and died at follow up from endocarditis complications.

The second study was a case report involving a 42 year old male with mitral PVI that had been done 4 years before aortic INVE^18^. Comorbidites included aortic valve regurgitation, rhemautic mitral disease, and mitral stenosis. He had undergone a St. Jude procedure of the mitral valve. The prosthesis type was unknown as was the source of INVE. The pathogen involved was Corynebacterium diphtheriae and he was treated with combined antibiotics + surgery. No complications were noted after aortic INVE and he was alive at follow up.

## Discussion

This review study showcases that INVE has been reported and well studied in native mitral valves after TAVI. While INVE in non mitral locations and INVE subsequent to non TAVI prostheses have also been reported in literature, these groups have not been studied as extensively. Given this discrepancy, it is not surprising that the INVE associations and outcomes here reflect what has been previously reported on mitral INVE after TAVI^1^. The cumulative findings do show that comorbidities such as DM and COPD were more closely associated with INVE than TAVIE. Based on these findings, it is plausible to say that these comorbidities may present a higher risk for mitral IE due to their high propensity for cardiopulmonary complications^25^. However the findings could be coincidental and need to be validated by using a larger INVE population and ruling out confounders based on raw data. On the other hand, the weaker correlation with CKD is consistent with data showing that TAVIE is closely associated with CKD^5^. This correlation may explain why renal failure was a significant cause of mortality in the TAVIE group. However, the number of patients with TAVIE and renal failure who initially had CKD was not available for analysis in our study and the true extent of this association would need to be explored further as well. Among other categories, the use of self expanding Corevalve TAVIs and the resulting mitral INVE was consistent with prior study findings which demonstrated the higher likelihood of mitral IE compared to TAVIE with the use of Corevalves^1^. As TAVIE presents a higher likelihood of serious complications early on, the study supports the expected finding that mortalities were higher in hospital compared with INVE. The converse finding of higher INVE mortality at follow up could be explained on the basis that conservative therapy may be less effective over time when the initial infection has had opportunity to spread systemically and lead to complications. Moreover, TAVIE is treated more aggressively with surgery on initial presentation and patients that survive may be less likely to have long term complications as compared to conservatively managed INVE patients. Appropriate management of INVE is therefore a subject of future study.

Regarding non mitral INVE after TAVI and mitral INVE after non TAVI, more investigations are needed. This review identified TAVI INVE cases that occurred as right heart INVE but the size of this group was insufficient for a subgroup analysis. Moreover, the number of SAVI INVE cases was even more limited along with the data on this group. The incidence of these cases does demonstrate however that there are alternate mechanisms for mitral and non mitral INVE in the setting of TAVIs, SAVIs, and other PVIs. Moreover, pathogens also seem to play a role in INVE as was suggested in studies showing that E coli can selectively target native mitral valves in the presence of prosthetic valves^20^.

Thus far, very few reviews have holistically examined the drivers and outcomes of INVE. Such reviews would have to identify many patients with this rare form of IE and acquire data on each individual patient, a process that would be extremely time consuming. On the other hand, smaller studies can more feasibly acquire individual patient data on categories of interest, but do have not enough power to make significant associations. Our review is the first to attempt a comprehensive analysis of this unusual phenomenon. It preliminarily suggests that the development of INVE after PVI is closely linked to comorbidity type, PVI type, procedural aspects of PVI, and pathogen type. It also indicates that the prognosis for INVE correlates with the presence of heart failure, renal failure, and septic shock. The significance of surgical vs medical management in long term survival is unclear and warrants future investigation.

## Limitations

There were some notable limitations in our study. First, only 16 studies fit the criteria for inclusion. Among TAVI studies, data was frequently acquired from the IE After TAVI Registry which after removal of duplicates limited the size of the TAVI INVE population. Moreover, the size of the SAVI INVE population was extremely small and this prevented any type of analysis on that subgroup. Overall, the discrepancies in these groups created the impression that INVE is a TAVI associated phenomenon when in reality data is lacking for a true comparison of the different PVIs. To this end, establishing the incidence, associations, and predictors for INVE requires more data and a larger population of INVE patients.

A second limitation in our analysis was the challenge of accounting for confounders. Although there were no concurrent infections and antibiotic prophylaxis was similar in TAVIE and TAVI INVE groups, performing a stratified analysis was challenging without data to compare the categories of interest. This could potentially change the significance of the associations seen in the study and would need to addressed using more raw data.

Another limitation involved the nature of the included studies themselves. One study was a multicenter review made up of a cohort that represented several international patient populations. All other studies drew data from unique hospital centers, each representing different patient populations. The final analysis required making a generalization about the associations in different patient populations and these associations may be different if comparable populations had been available for study.

Lastly, our study could not determine if studies excluded during full text review had unreported instances of INVE. As PVE is more common and studied more extensively than INVE, unreported INVE data could strengthen or weaken the associations in our study. Future research should be aimed towards acquiring more explicit data on INVE from studies that have reported on the phenomenon as well as studies that have not reported on the phenomenon.

## Conclusion

INVE is a complication frequently observed after transfemoral TAVIs among older patients. Diabetic and COPD patients appear to have a higher risk of developing this rare form of valvular IE, notably on mitral valves. Gram positive skin pathogens are frequently involved in INVE. Mortality in INVE is more common in those who develop renal failure, heart failure, and septic shock, particularly during long term follow up. The incidences, contributors, associations, and outcomes of INVE after SAVI and non aortic PVI are less well established and warrant future study.

## Abbreviations

IE: infective endocarditis
INVE: isolated native valve endocarditis
PVE: prosthetic valve endocarditis
NVE: native valve endocarditis
PVI: prosthetic valve implant
TAVI: transcatheter aortic valve implantation
TAVIE: transcatheter aortic valve implant endocarditis
SAVI: surgical aortic valve implantation
SAVIE: surgical aortic valve implant endocarditis
DM: diabetes mellitus
COPD: chronic obstructive pulmonary disease
CKD: chronic kidney disease

## Data Availability

All data produced in the present work are contained in the manuscript

